# Viral dispersion in open air

**DOI:** 10.1101/2021.01.18.21249463

**Authors:** Gabriella Trombini Machado, Claudia Ramos de Carvalho Pinto, Luisa Andrea Villanueva da Fonseca, Taissa Cristina dos Santos Ramos, Tuanny Fernanda Pereira Paggi, Beny Spira

## Abstract

The SARS-CoV-2 pandemic has revived the debate about the routes of virus transmission and their likelihoods. It is of utmost importance to assess the risks of contamination of susceptible people by infectious individuals and to evaluate the level of viral transmission in the community. Most countries have imposed non-pharmaceutical measures to contain SARS-CoV-2 transmission, including social distancing and mask wearing. Here we evaluated the spreading of viruses in open air using harmless *Escherichia coli* bacteriophages as a surrogate. Phages were sprayed towards Petri dishes seeded with bacteria at different lengths and angles. Median droplets size was 127 µm, similar to those produced by sneeze. Our results showed that the transmission rate decreased exponentially with distance. The highest recorded transmission rate was 9 × 10^−6^ PFU/plate when phages were sprayed from a 1 m distance, suggesting that the probability of transmission of a single virus at a 1 m distance is 1:100,000. These results agree with the WHO recommendation that face mask protection in an uncrowded well-ventilated space is not required.

## 1 Introduction

The current COVID-19 pandemic has renewed the discussion about the mechanisms through which respiratory viruses, such as SARS-CoV-2 spread and contaminate susceptible people. It is widely believed that SARS-CoV-2 spreads through droplets transmitted by infected people. These droplets, often released via coughing, sneezing or by regular breathing or talking (Riediker and Tsai, 2020), can be broadly classified into large (> 5 µm in diameter) and small droplets (≃5 µm in diameter) (Bar-On et al., 2020). Large droplets fall rapidly to the ground or to the nearest flat surface, while small droplets can remain airborne for an extended period of time (Wang et al., 2020). SARS-CoV-2 diameter is about 100 nm while the diameter of a droplet nuclei produced by coughing is about ten times larger (Yang et al., 2007). The mean size of droplets produced by sneezing varies substantially, but it is in the range of ten to hundreds of micrometers (Han et al., 2013).Viral transmission can occur by touching the nose or mouth mucosae with hands that have got previously in contact with surfaces contaminated by large droplets. Indeed, influenza control policies were based on the premise that most respiratory infections are transmitted by large respiratory droplets (Fennelly, 2020). However, more recent evidences has shown that humans produce infectious influenza droplets containing both aerosols with small particles and larger droplets (Fabian et al., 2008; Lindsley et al., 2010; Bischoff et al., 2013). The situation with SARS-CoV-2 is less certain, but it seems that COVID-19 propagates both via airborne and droplet transmission (Domingo et al., 2020; Morawska and Cao, 2020; Wilson et al., 2020; Tang et al., 2020). While contamination by large droplets should be directly proportional to the proximity to the index subject (Brankston et al., 2007), airborne transmission which occurs via small nuclei droplets in infectious aerosols has a much further reach. In confined spaces and particularly in superspreading events, such as choir rehearsals (Hamner, 2020) and prisons (Wallace, 2020) virus particles are released by infected people and accumulate in the air resulting in high transmission rates.

During the covid-19 pandemic governments and health agencies have been recommending or imposing different measures aiming at reducing virus transmission. Among them the requirement of social distancing and the use of face masks. The social distancing measures varied from country to country, from a recommendation to maintain 1-2 meters distance among each other to strict home confinement. The 1-2 meters distancing recommendation was based on the idea that virus transmission occurs via large droplets, as proximity to the index case was associated with higher transmission (Fennelly, 2020). Different countries adopted distinct approaches regarding face masks, while some countries have not recommended mask wearing at all, others have made face masks mandatory in all public spaces, indoors and outdoors, and others only in confined spaces such as public transportation and shops. As of January 2021, only a handful of countries do not recommend the use of masks under any circumstance, while most required the use of masks at least in places where social distancing is not possible (masks4all, 2020). In the most extreme cases, mask wearing was required everywhere, even outdoors in sparsely crowded spaces, such as parks and beaches. In some countries, only certain states or provinces require the use of masks as is the case of Australia, Brazil and the United States. In the state of São Paulo in Brazil, citizens older than 3-years old are required to wear masks in all public spaces, including in open air, under the threat of heavy fines (ALESP, 2020). Currently, the WHO recommends the use of face masks in settings where it is not possible to maintain a distance of at least 1 meter from others (WHO, 2020).

In principle, mask wearing by infected individuals may prevent the spread of viruses via large droplets, but only well fit respirators may offer protection from inhalation of infectious aerosols (Gawn et al., 2008). Nevertheless, randomised controlled trials in community sets have shown moderate to null protective effect of surgical masks or respirators on the transmission of respiratory viruses (Suess et al., 2012; Aiello et al., 2010, 2012; Cowling et al., 2009; Larson et al., 2010; MacIntyre et al., 2009; Simmerman et al., 2011; Bundgaard et al., 2020). Interestingly, there are no studies that evaluated the benefits of mask wearing in outdoors. Here we evaluated the spreading of viruses outdoors using *Escherichia coli* λ bacteriophages as a proxy for human respiratory viruses such as SARS-CoV-2. λ and other bacterial phages are similar in size to SARS-CoV-2 and have been used extensively in many fields of biology research, including as models for the study of airborne and not airborne eukaryotic viruses (Turgeon et al., 2014; Black et al., 2010; Kormuth et al., 2018). In addition, they are easy to manipulate and are entirely harmless to humans and to the environment. A set of experiments were conducted to test the effect of distance and movement relative to the infectious source in open settings.

## 2 Material and methods

### Strains and growth conditions

*E. coli* strain CSH109 (*ara* Δ*(gpt-lac)5 supE gyrA argE*_am_ *metB rpoB*) (Miller, 1992) was used as a λ Y1 phage host. Lysogeny broth (LB) and Lysogeny agar (L-agar) are the standard complex media used in this study (Miller, 1992). Bacteria were grown overnight in LB medium. On the next day, 0.1 ml of the bacterial culture were diluted in 4 ml R-Top-agar (Miller, 1992) and plated on L-agar. Bacteria were grown either in liquid or solid media at 37°C.

### Phage lysate preparation

λ phage lysate was prepared as described (Miller, 1992). Briefly, an overnight culture of strain CSH110 was diluted in LB medium and grown up to an OD_600_ of 0.3. 200 µl of this culture were mixed with 100 µl of a solution containing 10^7^ λ Y1 phages and 50 µl 1 M MgSO_4_. The mixture was incubated in a water bath at 37°C for 10 minutes. 5 ml R top-agar was then added and the mixture was poured onto an L-agar plate. The plates were incubated overnight. On the next day, the R Top-agar was scraped from the plate, which was subsequently washed with 1 ml LB medium supplemented with 0.01 M MgSO_4_ and the suspension was centrifuged at 5000 *xg* for 20 minutes. To the supernatant a 1/20 volume of chloroform was added. The lysate was stored under refrigeration until used.

Before every phage dispersion experiment the phage titer was evaluated by mixing serial dilutions of the lysate in SM solution (5.8 g NaCl, 2.0 g MgSO_4_, 50 ml Tris-HCl 1M pH 7,5 and 0.01% (m/v) gelatine in 1 L of water) with 5 ml R Top-agar and 100 µl of an overnight culture of strain CSH110. The mixture was poured over L-agar and the plates were incubated overnight at 37°C. Plaques were counted to obtain the PFU/ml concentration.

### Phage dispersion

The experiments that evaluated phage dispersion were performed using a trigger sprayer (Pulverizador Uniluk 500 ml, SP-Brazil) filled with a suspension containing a known concentration of phage λ Y1 diluted in a 0.1 M MgSO_4_. In most experiments, phage concentration in the spray bottle was ∼ 1.5 × 10^5^ PFU/ml. Each new phage dilution was tittered as described in the Phage lysate preparation section 2 to check the actual PFU concentration. A typical spraying event released a range of droplet sizes with a total volume of 0.7 ml. With the exception of one experiment, all others were carried out in the outdoors. The spray bottle was always handled in an upright position. Plates seeded with bacteria were held at different relative positions as shown in Figure S1 normally at 1, 2 or 3 meters away from the sprayer. The plates were kept open for 1 minute following the spraying and then incubated overnight at 37°C. On the next day, the plates were inspected for the presence of plaques and the PFU were counted.

### Transmission rate calculation

transmission rates were computed assuming a Poisson distribution of phage plaques and by counting the number of plates containing 0 PFU in each tier (Luria and Delbruck, 1943). The average number of plaques formation was calculated using the formula λ = −lnP_0_, where P_0_ represents the ratio of plates with no plaques over the total number of plates. To obtain the transmission rate the λ value was divided by 10^5^ (number of phages released by each spraying event).

### Droplets size measurement

Droplet size evaluation was performed at the LAPAR, Departamento de Ciências da Produção Agrícola (UNESP - Campus de Jaboticabal) using a Laser Diffraction Particle Size Analyzer (Mastersizer S^®^, version 2.19) as described (Fernandes et al., 2007). The spraying was driven by a trigger pump bottle (Pulverizador Uniluk 500 ml) filled either with water or with a 1 M NaCl solution, positioned at 40 cm away from the laser beam. No significant difference between water and NaCl droplets was observed. Droplet samples were evaluated at 1.5 s while a full spray jet passed through the laser. From the information generated by the Mastersizer software, the volumetric median diameter (VMD), the coefficient of uniformity (Span) and the percentage of droplet volumes were recorded.

## Statistical analysis

The standard error of the mean was calculated according to the formula 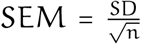 where SD is the standard deviation (Cumming et al., 2007). The coefficient of variation of the mean was calculated as follows: 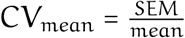.

## 3 Results

To evaluate the pattern of virus dispersion in open air, λ phages were sprinkled using a trigger spray bottle. Petri dishes seeded with λ-sensitive *E. coli* K-12 bacteria (strain CSH110) were used to detect the presence of phages in the sprayed emissions. To establish a correlation between the distance from the phage source and the number of PFU on the plates, the phage sprayer was positioned at the same height but at increasing 10 cm intervals from the plate (Figure 1B). In this experiment, the sprayer was filled with a suspension containing an average phage concentration of 3,575 PFU/ml. Each spray released 0.7 ml, i.e., 2,500 PFU. As expected, the farthest the plate from the bottle nozzle the lowest the number of plaques Figure 1B. When sprayed from 10 – 30 cm the phage plaques formed countless aggregates on the plate lawn, but from 40 to 100 cm the PFU number decreased exponentially, such that only 0.15% of the phages landed on the plates when sprayed from a 100 cm distance. Thus the ability of the phage droplets to travel through air in a straight line decreases exponentially with distance. This experiment was conducted indoors.

**Fig 1.**
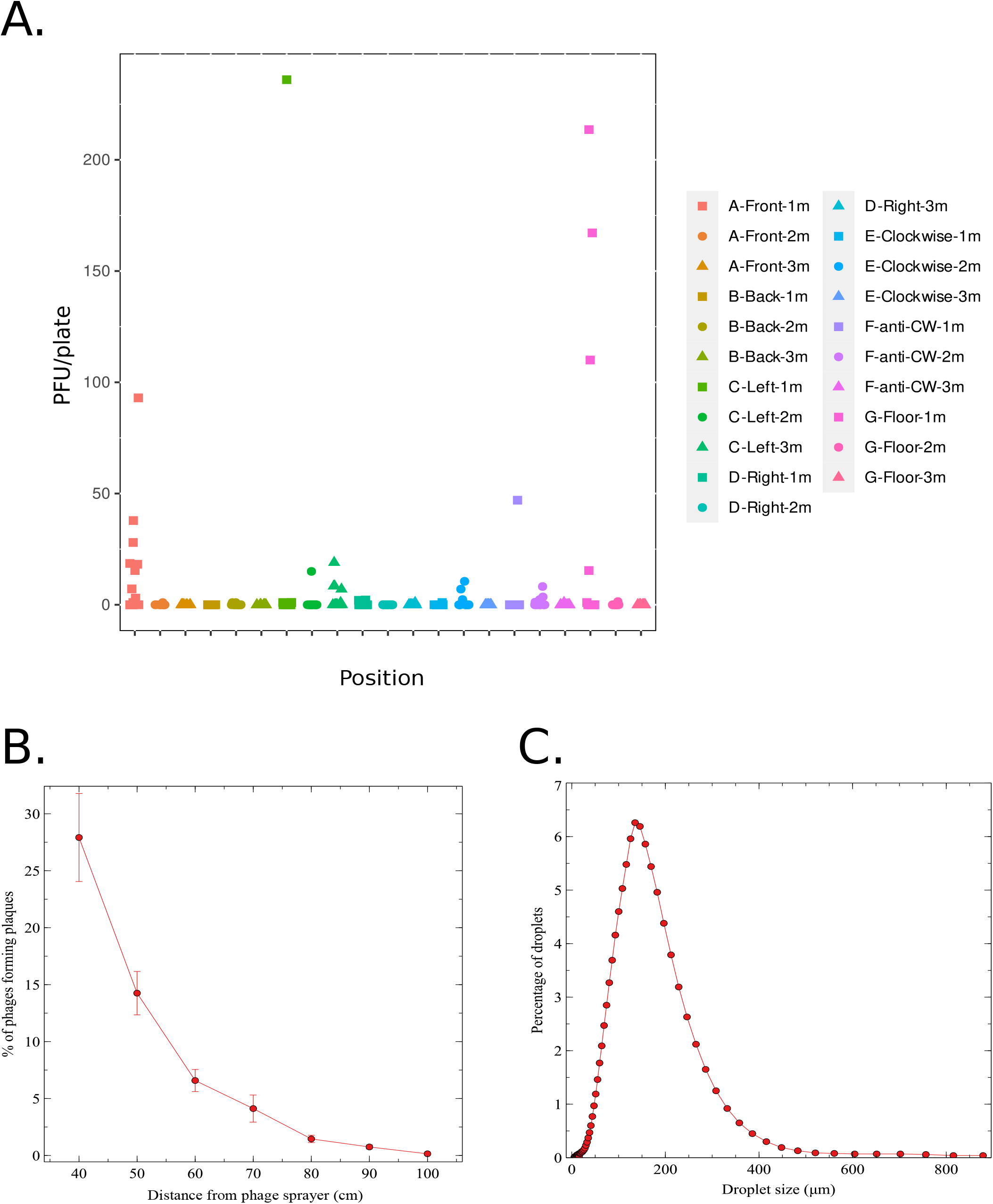
A. Distribution of PFU on plates sprayed with 10^5^ phages. Plates freshly seeded with *E. coli* cells were sprayed with λ phages as described in Figure S1A-C. Each time droplets containing approximately 10^5^ phages were released through the nozzle of a trigger sprayer. The plates containing the bacteria were kept open for 60 s following the spray and incubated overnight. On the next day the phage plaques were counted. The legend at the right describes the position of the plates relative to the phage sprayer. A1, A2 and A3 represent plates positioned at 1 m, 2 m and 3 m in front of the sprayer, respectively; B1, B2 and B3 represent plates positioned at the rear of the sprayer (at a 180° angle); C1, C2 and C3 represent plates positioned at the left with a 45° angle from the sprayer nozzle; D1, D2 and D3 is as above except that the plates were positioned at the right; E1, E2 and E3 represent plates moving from one side of the sprayer to the other making a 180° arch in a clockwise direction; F1, F2 and F3 is the same as above except that the plates were moved in an anti-clockwise direction. Finally, G1, G2 and G3 represent plates placed on the floor face up. With the exception of the G1, G2 and G3 setting, in all other experiments the plates were kept open in an upright position. In all cases, 1, 2 and 3 respectively represent 1 m, 2 m and 3 m distance between the sprayer and the open plate. B. PFU number in plates exposed to phage spraying at a 40-100 cm range. λ phages were sprayed towards plates seeded with *E. coli* from a spray bottle containing an average of 3,575 PFU/ml. The plates were positioned in front of the bottle nozzle 40 to 100 cm distance, as shown in Figure S1D. Each point represents the mean ±S.E.M. of at least 5 independent experiments. C. Distribution of droplets size emitted by the trigger sprayer. The size of the droplets emitted by a single discharge was assessed by a laser diffraction particle size analysis.

To test the pattern of phage dispersion in open air, bacteria-seeded plates were placed at three different distances (1, 2 or 3 m) and angles (0°, 45° or 180°) away from the sprayer. Figure S1A-C illustrates the several experimental settings that were conducted, all performed in open air. With the exception of one experiment, in which the plates were positioned on the floor facing upwards (Figure S1C), in all experimental sets the plates were handled upright (mimicking the position and angle of a human face), roughly 160 cm above ground and kept opened for 60 s following the spraying of phages. In these experiments, ∼ 10^5^ PFU were sprayed each time from a bottled containing a phage concentration of ∼ 5 × 10^5^ PFU/ml. Overall, phage plaques were observed in 42 out of 258 plates. The PFU/plate concentration obtained in each experiment is depicted in Figure 1A. The highest PFU count was observed in 3 plates that were placed at 1 m distance face up on the floor (299, 234 and 154 PFU), in one plate positioned at a 1 m distance 45° to the left (236 PFU) and in one plate placed at 1 m in front of the phage sprayer (93 PFU). However, even at the shortest distance (1 m) the majority of plates (63 out of 86) displayed no plaques at all. Table S1 shows the PFU in each plate that did present plaques. Most plates that were placed in front of the sprayer at a 1 meter distance (9 out of 12) developed at least one plaque, while 2 m distance, only one plate (out of 12) with a single plaque was observed. No plaques were found in plates distant 3 m from the sprayer. Under all other arrangements - back, right, left, clockwise or anti-clockwise movement, only the minority of plates presented plaques and at low numbers. One exception was a plate placed at 45° left at 1 m distance, which displayed 236 PFU. It is worth noticing that in some cases the droplets were carried by a weak wind gust blown at the precise time of spraying. The occurrence of wind gusts might explain the high number of plaques in some plates and the presence of PFUs in plates placed at distances larger than 1 meter. In the absence of wind (most plates placed more than 1 m away) recorded no plaques. Given the fact that the distribution of phages on the plates are rare positive events that occur in a continuum of space and time, the rate of transmission could be calculated by applying a Poisson distribution to the PFU results. The rate of transmission was thus computed for each experimental setting (Table 1). The highest transmission rate −9 × 10^−6^ was, as expected, observed in plates directly sprayed with phages from the short distance of 1 m. Under the other experimental conditions the transmission rate went from 0 to 5.4 × 10^−6^/plate. In principle, the transmission rate should have been directly proportional to the distance from the phage source, but this was not always the case. For instance, spraying from a 1 m or 3 m distance in the opposite direction of the plates (‘Back’ in Table 1) resulted in a null transmission rate, but at a 2 m distance, the transmission rate was 1.7 × 10^−6^. This divergence was caused by the incidence of weak wind gusts during some of the spraying events that shifted the path of the phage droplets towards or away from the plates as already pointed out.

**Table 1.** Transmission rates in each experimental setting.

| Position | Transmission rate | Position | Transmission rate |
| --- | --- | --- | --- |
| Front-1m | $9 \times 10^{-6}$ | Left-1m | $4.0 \times 10^{-6}$ |
| Front-2m | $8.7 \times 10^{-7}$ | Left-2m | $8.0 \times 10^{-7}$ |
| Front-3m | 0 | Left-3m | $4.4 \times 10^{-6}$ |
| Back-1m | 0 | Right-1m | $2.9 \times 10^{-6}$ |
| Back-2m | $1.7 \times 10^{-6}$ | Right-2m | 0 |
| Back-3m | 0 | Right-3m | $8.7 \times 10^{-7}$ |
| CW-1m | $8.7 \times 10^{-7}$ | anti-CW-1m | $8.7 \times 10^{-7}$ |
| CW-2m | $2.9 \times 10^{-6}$ | anti-CW-2m | $4.0 \times 10^{-7}$ |
| CW-3m | 0 | anti-CW -3m | $8.7 \times 10^{-7}$ |
| Floor-1m | $5.4 \times 10^{-6}$ | | |
| Floor-2m | $8.7 \times 10^{-7}$ | | |
| Floor-3m | 0 |  |  |

Virus particles can be released by infected individuals through droplets having a range of sizes, from less than 1 µm to more than 100 µm (Han et al., 2013; Dhand and Li, 2020; Riediker and Tsai, 2020). To assess the size of the droplets produced by our sprayer, a laser diffraction particle size analysis was conducted. The size of the produced droplets ranged from 5 µm to 815 µm. The median size was 127 µm, with 50% of the droplets between 60 and 160 µm. Figure 1C show the distribution of droplet sizes discharged by the sprayer used in this study.

## 4 Discussion

In this study we followed the spreading of phages outdoors as a surrogate for virus transmission in public open spaces. The experiment was purposely set in open air where wind and other physical conditions might have some influence on the transmission rates. At the closest distance and straight angle from the phage spreading source (1 m Front in Figure 1A) the calculated transmission rate was slightly less than 10^−5^/plate, suggesting that even at this close distance (akin to an infectious individual sneezing towards a susceptible subject), the probability of getting infected with a single phage was 1:100,000.

Ever since the pioneer work of Ellis and Delbrück (1939) on the infection and growth of bacterial viruses, it is known that bacteriophage adsorption to the bacterial host stops upon dilution of the bacteria:phage mixture. Namely, these authors showed that by simply diluting the phage-bacteria mixture 10, 000× the infection is effectively brought to a halt. Phage dispersion in open air follows a similar dilution path, and it is even more pronounced than in liquid media because only small droplets or aerosols continue floating in the air, while large size phage-containing droplets rapidly fall to the ground. In addition, in open air the extent of dilution is orders of magnitude higher than 10,000 times. Thus, unlike a non-ventilated confined space where small virus particles accumulate in the air for an indefinite amount of time or until reaching a flat surface, in the outdoors these airborne particles simply disperse or get diluted. Accordingly, it has been shown that outdoor air in residential and urban areas was mostly devoid of SARS-CoV-2 particles in both northern and southern Italy during the peak of the pandemic in May 2020 (Chirizzi et al., 2020).

The amount of SARS-CoV-2 particles emitted through cough varies from 0.000277 to 36,030/cm^3^/cough (Riediker and Tsai, 2020). A typical virus emitter coughs 250 ml of 0.277 virus particles/cm^3^ (Riediker and Tsai, 2020), i.e., 69.25 copies of virus per cough. While a single cough releases ∼3,000 droplets, sneezing might produce an estimate of 40,000 droplets (Cole and Cook, 1998). However, no reliable estimates of SARS-CoV-2 infectious particles released by sneezing could be found. Each phage spraying event emitted ∼0.7 ml, containing approximately 10^5^ active phages. This number is thus about 1,444 times higher than the average number of SARS-CoV-2 particles released in a single cough by a typical emitter. The precise infectious dose for SARS-CoV-2 is unknown, but it is believed to be closer to that of SARS and lower than that for MERS - requiring something between 500 and 50,000 PFU (Ryan et al., 2020). The phage dispersion experiments showed the maximal transmission rate of 9 × 10^−6^, which indicates that only 1 in ∼100, 000 phage particles are able to reach the plate at a 1 m distance. At larger distances the transmission rate was even lower. A spray at a 1 m distance is akin to a sneeze or cough directed towards the face of a nearby individual. In that case, our results suggest that an infectious individual coughing or sneezing should shed 10^7^ virus particles in order to infect a susceptible person at a 1 m distance with a minimal infectious dose of 100 virus particles, provided that the coughing/sneezing is an one-time event in open air or in an well-ventilated space.

Can SARS-CoV-2 and phage particles be compared? Phages have been used as surrogates for eukaryotic viruses in many instances (Turgeon et al., 2014; Black et al., 2010; Kormuth et al., 2018). In terms of size, the circular SARS-CoV-2 has a diameter of 100 nm, while the λ phage has an icosahedral head with about 50-60 nm in diameter, and a tail with about 150 nm in length (Abedon, 2005; Furth et al., 1983). Given their similar sizes both types of particles should follow comparable spreading paths. In addition, our sprayer produced many relatively large droplets (Figure 1C) that would rapidly fall on the ground. Indeed, the presence of phages in heavy droplets could be inferred from the observation of PFU in the experiments in which phages were sprayed in the direction of plates placed face up on the ground. It is likely that SARS-CoV-2 in these large droplets would follow the same pathway as λ phages did. The median diameter size of the droplets produced by our sprayer was 127 µm, which is compatible with sneeze droplets size but larger than that of cough particles (Han et al., 2013). A droplet diameter of 127 µm corresponds to a volume of 0.001 µl. Given that the phage concentration in the sprinkler was 150,000 phages/ml, each droplet should carry about 0.15 phages, i.e., roughly only 1 in every 6 droplets contained a phage particle. We can therefore be confident that each observed plaque on the plates derives from a single phage. The reported size of the droplets produced by human respiratory activities is inconsistent (Han et al., 2013; Fabian et al., 2008; Yang et al., 2007; Buckland and Tyrrel, 1964). Nevertheless, the diameter of droplets produced by cough or speech tend to be smaller than the ones produced by our sprayer. Aerosols, in particular, consist of very small particles (< 5 µm) that can in the absence of air currents or ventilation remain airborne for extended periods of time (Fennelly, 2020). However, in open air, these small droplets tend to dilute immediately as already pointed out. The ones that may be the cause of concern in the open air are the high-velocity larger droplets produced by sneeze. Here we showed that this type of infectious droplets are able to infect an open Petri dish with a 9 cm diameter (area = 64 cm^2^) at a 1 m distance at the low rate of 10^−5^. At a 2 m distance the transmission rate drops even further. It should be noticed that the exposed area in a Petri dish is considerably larger than the total mucosal area in a human face.

In conclusion, we presented here a simplified test for the dispersion of viruses in open air. For obvious reasons, such an experiment could not be performed with live human viruses outside the laboratory, but by using a bacterial virus that is completely innocuous to all living beings, except for a narrow range of *E. coli* strains, harmless to the environment and that resembles in size respiratory viruses it was possible to evaluate the pattern of outdoors transmission. Our results suggest that virus transmission at a 1 m distance, i.e. not very crowded open air settings, such as streets and parks or in well-ventilated sites has a very small likelihood. Therefore, the use of face masks or any other protective shield outdoors is not required as long as a minimal distance of 1 m is kept among each other, as already recommended by the WHO and adopted by many countries (WHO, 2020; masks4all, 2020).

## Data Availability

The authors confirm that the data supporting the findings of this study are available within the article

## Acknowledgements

We are grateful to Fundação de Amparo à Pesquisa do Estado de São Paulo (FAPESP). B.S. is a recipient of a productivity scholarship from the Conselho Nacional de Desenvolvimento Científico e Tecnológico (CNPq). C.R.C.P, G.T.M., L.A.V.F. T.C.S.R. and T.F.P.P. are recipients of CAPES graduate scholarships. The authors thank Dr. Marcelo da Costa Ferreira for kindly performing the analysis of droplets size.

## Supporting Information

**Table S1.** Individual plates with positive number of plaques.

|  |  |  |  |
| --- | --- | --- | --- |
| Front-1m | 93 | Left-1m | 236 |
| Front-1m | 1 | Left-1m | 1 |
| Front-1m | 3 | Left-1m | 1 |
| Front-1m | 53 | Left-1m | 1 |
| Front-1m | 10 | Left-2m | 15 |
| Front-1m | 26 | Left-3m | 19 |
| Front-1m | 11 | Left-3m | 1 |
| Front-1m | 20 | Left-3m | 5 |
| Front-1m | 13 | Left-3m | 6 |
| Front-2m | 1 | Left-3m | 6 |
| Back-2m | 1 | CW-1m | 1 |
| Back-2m | 1 | anti-CW-1m | 47 |
| Right-1m | 2 | Floor-1m | 1 |
| Right-1m | 1 | Floor-1m | 234 |
| Right-1m | 3 | Floor-1m | 154 |
| Right-3m | 1 | Floor-1m | 299 |
|  |  | Floor-1m | 11 |
|  |  | Floor-2m | 2 |

**Fig S1.**
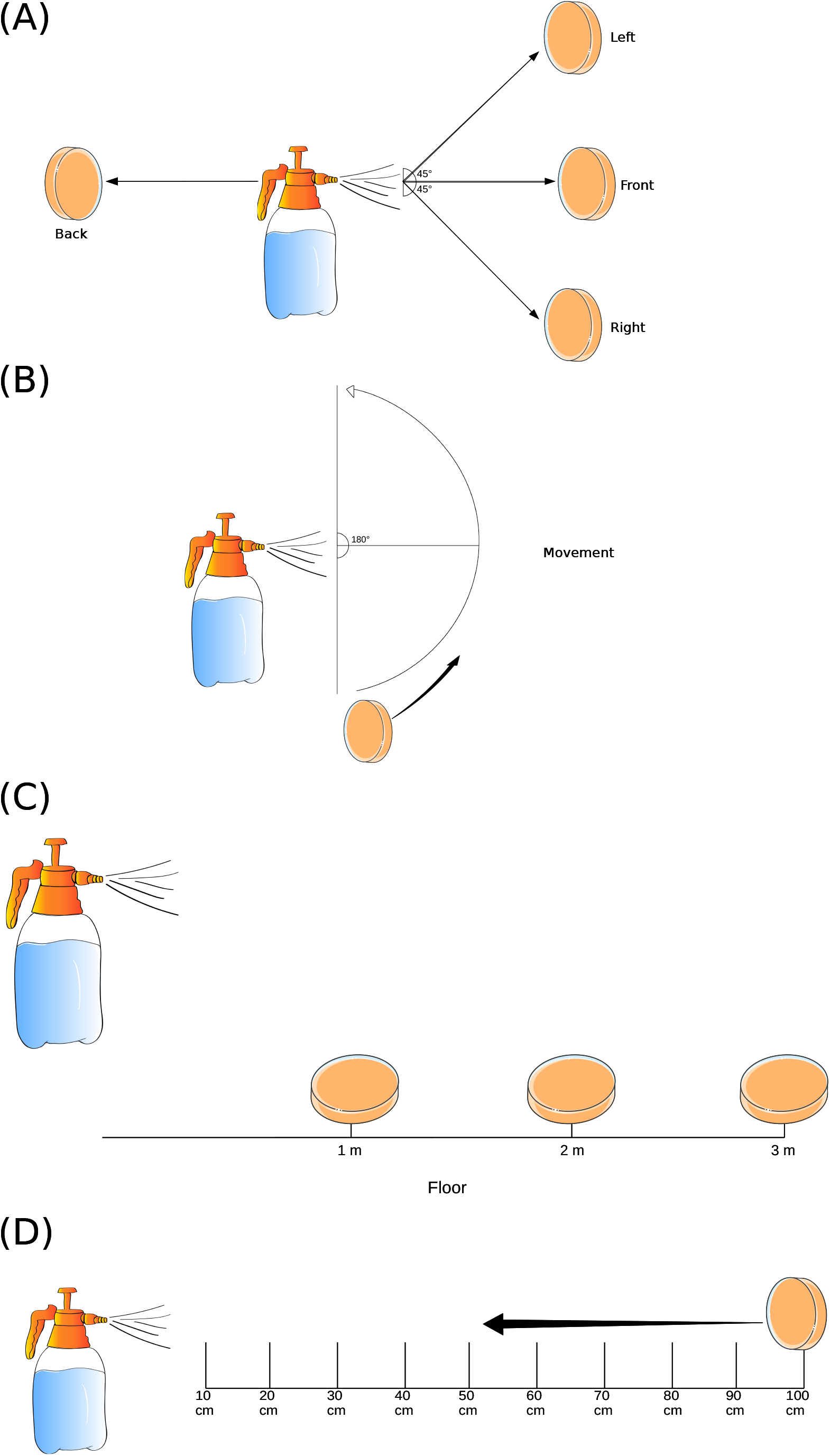
Experimental settings of virus spreading. A trigger sprayer containing ∼ 1.5 · 10^5^ PFU/ml of λ phages was used to spray top-agar plates freshly seeded with ∼ 5 · 10^8^ susceptible *E. coli* bacteria. Each spray (0.7 ml) contained approximately 10^5^ phages. (A) Phages were sprayed towards plates positioned at the same height but at different horizontal arrangements: Front, Back, 45° to the left and 45° to the right. In each situation, the plates were positioned at 1, 2 or 3 m away from the phage source (sprayer). Plates lids were removed and the plates were held vertically for 1 minute following the spray shot. (B) immediately following the spraying event lidless plates held vertically at 1, 2 or 3 m from the phage source were circularly moved at walking pace either clockwise or anti-clockwise. (C) Open plates were held vertically 10-100 cm away from the sprayer. The plates were kept open for a few seconds following the spray event.

## References

Abedon, S.T., 2005. The bacteriophages. Oxford University Press.

Aiello, A.E., Murray, G.F., Perez, V., Coulborn, R.M., Davis, B.M., Uddin, M., Shay, D.K., Waterman, S.H., Monto, A.S., 2010. Mask use, hand hygiene, and seasonal influenza-like illness among young adults: a randomized intervention trial. The Journal of infectious diseases 201, 491–498. doi:10.1086/650396.

Aiello, A.E., Perez, V., Coulborn, R.M., Davis, B.M., Uddin, M., Monto, A.S., 2012. Facemasks, hand hygiene, and influenza among young adults: a randomized intervention trial. PloS one 7, e29744. doi:10.1371/journal.pone.0029744.

ALESP, 2020. Dispõe sobre o uso geral e obrigatório de máscaras de proteção facial no contexto da pandemia da covid-19 e dá medidas correlatas. URL: https://www.al.sp.gov.br/norma/193701.

Bar-On, Y.M., Flamholz, A., Phillips, R., Milo, R., 2020. Sars-cov-2 (covid-19) by the numbers. eLife 9. doi:10.7554/eLife.57309.

Bischoff, W.E., Swett, K., Leng, I., Peters, T.R., 2013. Exposure to influenza virus aerosols during routine patient care. The Journal of infectious diseases 207, 1037–1046. doi:10.1093/infdis/jis773.

Black, E., Cascarino, J., Guan, D., Kniel, K., Hicks, D., Pivarnik, L., Hoover, D., 2010. Coliphage as pressure surrogates for enteric viruses in foods. Innovative Food Science & Emerging Technologies 11, 239–244. doi:10.1016/j.ifset.2009.07.004.

Brankston, G., Gitterman, L., Hirji, Z., Lemieux, C., Gardam, M., 2007. Transmission of influenza a in human beings. The Lancet. Infectious diseases 7, 257–265. doi:10.1016/S1473-3099(07)70029-4.

Buckland, F.E., Tyrrel, D.A., 1964. Experiments on the spread of colds. 1. laboratory studies on the dispersal of nasal secretion. The Journal of hygiene 62, 365–377. doi:10.1017/s0022172400040080.

Bundgaard, H., Bundgaard, J.S., Raaschou-Pedersen, D.E.T., von Buchwald, C., Todsen, T., Norsk, J.B., Pries-Heje, M.M., Vissing, C.R., Nielsen, P.B., Winsløw, U.C., Fogh, K., Hasselbalch, R., Kristensen, J.H., Ringgaard, A., Porsborg Andersen, M., Goecke, N.B., Trebbien, R., Skovgaard, K., Benfield, T., Ullum, H., Torp-Pedersen, C., Iversen, K., 2020. Effectiveness of adding a mask recommendation to other public health measures to prevent sars-cov-2 infection in danish mask wearers : A randomized controlled trial. Annals of internal medicine doi:10.7326/M20-6817.

Chirizzi, D., Conte, M., Feltracco, M., Dinoi, A., Gregoris, E., Barbaro, E., La Bella, G., Ciccarese, G., La Salandra, G., Gambaro, A., et al., 2020. Sars-cov-2concentrations and virus-laden aerosol size distributions in outdoor air in north and south of italy. Environ Int 146, 106255.

Cole, E.C., Cook, C.E., 1998. Characterization of infectious aerosols in health care facilities: an aid to effective engineering controls and preventive strategies. American journal of infection control 26, 453–464. doi:10.1016/s0196-6553(98)70046-x.

Cowling, B.J., Chan, K.H., Fang, V.J., Cheng, C.K.Y., Fung, R.O.P., Wai, W., Sin, J., Seto, W.H., Yung, R., Chu, D.W.S., Chiu, B.C.F., Lee, P.W.Y., Chiu, M.C., Lee, H.C., Uyeki, T.M., Houck, P.M., Peiris, J.S.M., Leung, G.M., 2009. Facemasks and hand hygiene to prevent influenza transmission in households: a cluster randomized trial. Annals of internal medicine 151, 437–446. doi:10.7326/0003-4819-151-7-200910060-00142.

Cumming, G., Fidler, F., Vaux, D., 2007. Error bars in experimental biology. J Cell Biol 177, 7–11.

Dhand, R., Li, J., 2020. Coughs and sneezes: Their role in transmission of respiratory viral infections, including sars-cov-2. American journal of respiratory and critical care medicine 202, 651–659. doi:10.1164/rccm.202004-1263PP.

Domingo, J.L., Marquès, M., Rovira, J., 2020. Influence of airborne transmission of sars-cov-2 on covid-19 pandemic. a review. Environmental research 188, 109861. doi:10.1016/j.envres.2020.109861.

Ellis, E.L., Delbrück, M., 1939. The growth of bacteriophage. The Journal of general physiology 22, 365–384. doi:10.1085/jgp.22.3.365.

Fabian, P., McDevitt, J.J., DeHaan, W.H., Fung, R.O.P., Cowling, B.J., Chan, K.H., Leung, G.M., Milton, D.K., 2008. Influenza virus in human exhaled breath: an observational study. PloS one 3, e2691. doi:10.1371/journal.pone.0002691.

Fennelly, K.P., 2020. Particle sizes of infectious aerosols: implications for infection control. The Lancet. Respiratory medicine 8, 914–924. doi:10.1016/S2213-2600(20)30323-4.

Fernandes, A.P., Parreira, R.S., Ferreira, M.C., Romani, G.N., 2007. Caracterização do perfil de deposição e do diâmetro de gotas e otimização do espaçamento entre bicos na barra de pulverização. Engenharia Agrícola 27, 728–733.

Furth, M., Wickner, S., Hendrix, R., Roberts, J., Stahl, F., Weisberg, R., 1983. Lambda II. Cold Spring Harbor Laboratory Cold Spring Harbor, NY.

Gawn, J., Clayton, M., Makison, C., Crook, B., 2008. Evaluating the protection afforded by surgical masks against influenza bioaerosols: gross protection of surgical masks compared to filtering facepiece respirators. Health Safety Exec.

Hamner, L., 2020. High sars-cov-2 attack rate following exposure at a choir practice—skagit county, washington, march 2020. MMWR. Morbidity and Mortality Weekly Report 69.

Han, Z.Y., Weng, W.G., Huang, Q.Y., 2013. Characterizations of particle size distribution of the droplets exhaled by sneeze. J R Soc Interface 10, 20130560. doi:10.1098/rsif.2013.0560.

Kormuth, K.A., Lin, K., Prussin, A.J., Vejerano, E.P., Tiwari, A.J., Cox, S.S., Myerburg, M.M., Lakdawala, S.S., Marr, L.C., 2018. Influenza virus infectivity is retained in aerosols and droplets independent of relative humidity. The Journal of Infectious Diseases 218, 739–747. doi:10.1093/infdis/jiy221.

Larson, E.L., Ferng, Y.h., Wong-McLoughlin, J., Wang, S., Haber, M., Morse, S.S., 2010. Impact of non-pharmaceutical interventions on uris and influenza in crowded, urban households. Public health reports (Washington, D.C. : 1974) 125, 178–191. doi:10.1177/003335491012500206.

Lindsley, W.G., Blachere, F.M., Thewlis, R.E., Vishnu, A., Davis, K.A., Cao, G., Palmer, J.E., Clark, K.E., Fisher, M.A., Khakoo, R., Beezhold, D.H., 2010. Measurements of airborne influenza virus in aerosol particles from human coughs. PloS one 5, e15100. doi:10.1371/journal.pone.0015100.

Luria, S., Delbruck, M., 1943. Mutations of bacteria from virus sensitivity to virus resistance. Genetics 28, 491–511.

MacIntyre, C.R., Cauchemez, S., Dwyer, D.E., Seale, H., Cheung, P., Browne, G., Fasher, M., Wood, J., Gao, Z., Booy, R., Ferguson, N., 2009. Face mask use and control of respiratory virus transmission in households. Emerging infectious diseases 15, 233–241. doi:10.3201/eid1502.081167.

masks4all, 2020. What countries require masks in public or recommend masks? URL: https://masks4all.co/what-countries-require-masks-in-public/.

Miller, J.H., 1992. A Short Course In Bacterial Genetics: A Laboratory Manual And Handbook For Escherichia coli And Related Bacteria. Cold Spring Harbor Laboratory, Cold Spring Harbor, N.Y.

Morawska, L., Cao, J., 2020. Airborne transmission of sars-cov-2: The world should face the reality. Environment international 139, 105730. doi:10.1016/j.envint.2020.105730.

Riediker, M., Tsai, D.H., 2020. Estimation of viral aerosol emissions from simulated individuals with asymptomatic to moderate coronavirus disease 2019. JAMA network open 3, e2013807. doi:10.1001/jamanetworkopen.2020.13807.

Ryan, K.A., Bewley, K.R., Fotheringham, S.A., Brown, P., Hall, Y., Marriott, A.C., Tree, J.A., Allen, L., Aram, M.J., Brunt, E., et al., 2020. Dose-dependent response to infection with sars-cov-2 in the ferret model: evidence of protection to re-challenge. bioRxiv.

Simmerman, J.M., Suntarattiwong, P., Levy, J., Jarman, R.G., Kaewchana, S., Gibbons, R.V., Cowling, B.J., Sanasuttipun, W., Maloney, S.A., Uyeki, T.M., Kamimoto, L., Chotipitayasunondh, T., 2011. Findings from a household randomized controlled trial of hand washing and face masks to reduce influenza transmission in bangkok, thailand. Influenza and other respiratory viruses 5, 256–267. doi:10.1111/j.1750-2659.2011.00205.x.

Suess, T., Remschmidt, C., Schink, S.B., Schweiger, B., Nitsche, A., Schroeder, K., Doellinger, J., Milde, J., Haas, W., Koehler, I., Krause, G., Buchholz, U., 2012. The role of facemasks and hand hygiene in the prevention of influenza transmission in households: results from a cluster randomised trial; berlin, germany, 2009-2011. BMC infectious diseases 12, 26. doi:10.1186/1471-2334-12-26.

Tang, S., Mao, Y., Jones, R.M., Tan, Q., Ji, J.S., Li, N., Shen, J., Lv, Y., Pan, L., Ding, P., et al., 2020. Aerosol transmission of sars-cov-2? evidence, prevention and control. Environment international 144, 106039.

Turgeon, N., Toulouse, M.J., Martel, B., Moineau, S., Duchaine, C., 2014. Comparison of five bacteriophages as models for viral aerosol studies. Appl Environ Microbiol 80, 4242–4250. doi:10.1128/aem.00767-14.

Wallace, M., 2020. Covid-19 in correctional and detention facilities—united states, february–april 2020. MMWR. Morbidity and mortality weekly report 69.

Wang, Y., Xu, G., Huang, Y.W., 2020. Modeling the load of SARS-CoV-2 virus in human expelled particles during coughing and speaking. PLOS ONE 15, e0241539. doi:10.1371/journal.pone.0241539.

WHO, 2020. Coronavirus disease (covid-19): Masks. URL: https://www.who.int/emergencies/diseases/novel-coronavirus-2019/question-and-answers-hub/q-a-detail/coronavirus-disease-covid-19-masks.

Wilson, N., Corbett, S., Tovey, E., 2020. Airborne transmission of covid-19. Br Med J 370.

Yang, S., Lee, G.W.M., Chen, C.M., Wu, C.C., Yu, K.P., 2007. The size and concentration of droplets generated by coughing in human subjects. Journal of aerosol medicine : the official journal of the International Society for Aerosols in Medicine 20, 484–494. doi:10.1089/jam.2007.0610.

